# Influencing Factors for the Persistence of SARS-Cov-2 (Covid-19) exposed in Environmental Matrices and Disinfection Methods: Systematic Review

**DOI:** 10.1101/2022.06.15.22276331

**Authors:** Chaw Chaw Yu, Thein Hlaing, Kyaw Myo Tun

**Author notes:** **Corresponding author** Dr Chaw Chaw Yu, Yangon, Myanmar.

## Abstract

**Background:** Since the COVID-19 pandemic has been pestilential over a considerable duration, global deployment and financial crisis could not be reversed as before. It brought up essentials to allow the nations back to work with effective preventive measures. This review intended to evaluate the persistence of SARS-CoV-2(COVID-19) exposed in the environmental matrices, influencing factors on the virus persistence and disinfection methods.

**Methods:** Applying the PRISMA 2020 tool, MEDLINE/PubMed, HINARI, and Google Scholar were primarily explored. Data were extracted, entered into the modified data extraction forms and analysed narratively. Quality appraisal was done by the Mixed-Methods Appraisal Tool. The findings were presented descriptively.

**Results:** Persistence of SARS-CoV-2 was revealed <4 hours on aluminium, 4 hours on copper, 24 hours on cardboard, 44 hours on glass, 48 hours on stainless steel, 72 hours on plastic, 92 hours on polystyrene plastic, 1.1-1.2 hours in the air, 7 days (higher titer) to 3 days(lower titer) in wastewater. Virus decaying was noted 5-10 times faster at 27°C than at 10°C and 2-5 times faster with 65% relative humidity (RH) than with 40% and 100% RH. Virus infectivity was reduced by far-UVC-(222 nm) light for 90%-(8 minutes), 95%-(11 minutes), 99%-(16 minutes) and 99.99%-(25 minutes). Sodium hypochlorite (800 g/m^3^) and ammonium-based detergents were remarkably effective for preliminary disinfection.

**Conclusions:** This review identified the duration of SARS-CoV-2 survival in environmental matrices for both healthcare and non-healthcare settings. The study explored the impacts of environmental factors on the virus and effective disinfection methods to be considered accordingly to the findings.

## Introduction

Coronavirus disease 2019 (COVID-19) is a newly discovered infectious disease caused by a new human virus of the coronaviridae family which was firstly identified in Wuhan, the capital of Hubei province, China in December 2019. It was officially named COVID-19 by the World Health Organisation (WHO) on 12^th^ February 2020 and also named Severe Acute Respiratory Syndrome Coronavirus-2(SARS-CoV-2), because of its 88% genetic similarity with SARS-like coronaviruses of bat origin [1]. The first virus was discovered over 100 years ago and found the human infection in the late 1800s. Over 150 species of RNA virus were discovered and Coronavirus is one of them [2]. Viruses can be dispersed through aerosols from coughing, sneezing and talking, which in turn contaminate the environment. Its single droplet may easily contain an infectious dose [3].

Enveloped respiratory viruses, which are though more vulnerable to environmental stress than non-enveloped viruses, have been shown to persist on surfaces for a certain period. Enveloped respiratory viruses may persist on common hard surfaces longer and cause the potential risk of infection to whoever touches those contaminated surfaces [4]. When the expelled microorganisms persist with adequate doses of viruses for long enough in the environment to contact with other hosts, indirect and widespread contraction of disease occurs. Once the disease is transmitted from indirect contact, it is challenging to trace the disease’s origin, especially in case of contamination from pre-symptomatic patients [5]. While SARS-CoV-2 is assumed as droplet transmission by WHO claimed that viruses may be transmissible through aerosols and may also survive in water apart from contracting via contaminated surfaces [6]. SARS-CoV-2 can persist for a few hours in the air after the generation of aerosols [7]. Virus contamination on air exhaust outlets, means, viruses can be travelled by air [8]. Besides, SARS-CoV-2 RNA is detected in faeces suggesting virus replication and shedding through patients’ GI tract [9]. With the likelihood of virus survival in faeces and water, attention should be placed on water-related virus exposures as well [10]. Regarding fomites, these include high touch surfaces of porous and non-porous materials in both healthcare and non-healthcare settings. High touch surfaces of fomites are the highest risk of virus transmission through contaminated environments [11]. Various studies of SARS-CoV-2 persistence in common public spaces explored the virus survival duration on the surfaces of furniture, household fixed items, electronic objects, and stairway rails, floors, walls, shelves and countertops. According to WHO, SARS-CoV-2 can survive up to several hours on some porous surfaces such as cloths, cardboards and wood while the virus can be persisting up to several days on different porous materials such as the outer layer of a medical mask [12]. SARS-CoV-2 on non-porous surfaces like copper, glass and stainless steel may persist up to many hours whereas the virus can survive on plastic for many days. Foremost, healthcare settings are essentially under concern for contamination with microscopic virus particles exhaled from patients. Emergency departments, intensive care units, wards, primary health clinics, facilities used for isolation of COVID-19 patients, medical gadgets, surgical tools, instruments in operation theatres, rubbish & waste released from healthcare places (masks & gloves, etc.) are very risky of virus contamination and surface persistence unless effective disinfection measures [13]. The door handle, toilet bowl and sink were found to be the test positive for the virus. Airflows equipment like vents and Personal Protective Equipment (PPE) also resulted in positive virus tests [8]. Factors influencing the persistence of SARS-CoV-2 (Covid-19) in different environmental metrics/surfaces are vital to acknowledge in consideration of effective disinfection methods for the prevention of the disease. Many international and national guidelines for preventive measures were developed based on the available information for SARS-CoV-2 so far. The review was to explore the persistence of SARS-CoV-2(Covid-19) exposed in environmental matrices (air, water, faeces)/fomites surfaces (porous and non-porous) with influencing factors for the virus persistence in environments (such temperature, humidity, pH) and different methods of disinfection. The resulting information will support the relevant authorities in modification of Covid-19 preventive measures accordingly.

## Methods

A systematic review methodology following PRISMA guidelines and its checklist was adopted for this study (S1 Table). PICO (population, intervention, comparison, outcomes) guidelines were used to formulate the research question; “What are the influencing factors for the persistence of SARS-CoV-2(COVID-19) exposed in environmental matrices and disinfection methods?” SARS-CoV-2(COVID-19) in environments was assumed as a problem desired to examine (P). The exposure of SARS-CoV-2 with influencing factors (Temperature, Humidity, UV radiation & pH) and different disinfection methods was assumed as intervention (I). The different duration of SARS-CoV-2 persistence in different environmental matrices, various impacts of influencing factors on SARS-CoV-2 in environments and different disinfection methods were measured as outcomes (O). These PICO criteria became key terms for the literature search.

### Eligibility Criteria

When determining the type of studies, all primary studies published full-text in English since the beginning of 2020, which were conducted in both healthcare and non-healthcare settings were counted. Systematic, editorial and narrative reviews, government and organisation guidelines, patents, books and data linked with various commercial disinfection products were excluded. The consideration of healthcare settings included but was not limited to acute-care hospitals, long-term care facilities, nursing homes and skilled nursing facilities, physicians’ offices, urgent-care centres, outpatient clinics, home healthcare (i.e., care provided at home by professional healthcare providers), emergency medical services, mobile healthcare services and medical clinic embedded with a workplace or school. In non-healthcare settings, the review included the community facilities (schools/daycare centres/ community centres/businesses) and common public spaces (plazas/squares/parks/sidewalks/ streets). Any findings, even a single report of the aforesaid outcomes, such as various degrees of SARS-CoV-2 persistence in environments, influencing factors’ impacts on the virus persistence in environments and different methods of disinfection were presumed as eligible for inclusion (Table 1).

**Table.1.** Summary of Eligibility Criteria.

| Inclusion Criteria | Exclusion criteria |
| --- | --- |
| • Primary research studies<br>• Full text published as peer-reviewed journals or preprint papers with high quality or other high-quality papers determined by the review team<br>• SARS-CoV-2 (Covid-19) in environments<br>• Influencing factors for the persistence of SARS-CoV-2 (Covid-19) in environmental matrices<br>• Disinfection methods for SARS-CoV-2 (Covid-19)<br>• Published in the English language<br>• Articles published since the beginning of 2020 | • Secondary research such as systematic reviews, editorial reviews, narrative reviews, reports, letters, Government and organization guidelines, patents and books<br>• Another virus except for SARS-CoV-2<br>• Do not examine SARS-CoV-2 in biological matrices<br>• Other languages except for English<br>• Studies without access to full articles<br>• Articles published before 2020<br>• Data with commercial products of disinfection agents (e.g. studies for comparison of 2 commercial products) |

### Bibliographic Search

The prime databases used for searching articles were Google Scholar, HINARI and MEDLINE/PubMed. Other databases such as EBSCOHOST and Cochrane Library were also explored. By using the predefined keywords, pilot screening was performed and keywords were modified accordingly. Besides, the search strategy was tested with Boolean Operators by two independent investigators and compared the total number of eligible papers found. Whenever significant differences in the number of searched papers had occurred, thorough discussions between investigators were performed to optimise the keywords and searching mechanism. The potential title and abstract screening process was initiated after the confirmation of keywords and search strategy. The full-text screening was then proceeded to filter the eligible articles. Additionally, a manual search and screening of the reference lists were elaborated. Moreover, Medical Subject Headings (MeSH) terms and specific symbols such as the asterisk-(*) marks and dollar signs-($) were combined in the search mechanism to identify truncation or find the related terms to find relevant articles.

### Identification and Selection of Studies

The fundamental steps for studies’ identification and selection were the title and abstract screening, saving and sorting out potential articles in software like EndNote, filtering the duplicated papers, screening for eligibility of studies and compiling a final selection for analysis. The self-developed screening tool was used to check if the titles/abstracts/full-texts met any of the pre-defined eligibility criteria. Once the titles/abstracts/full-texts met the inclusion criteria or remained unclear, papers were saved for the next-step screening process otherwise excluded. PRISMA 2009 flow diagram was utilised for the screening process. The selection and revision process of obtained papers were performed by two independent reviewers under the supervision of the immediate supervisor.

### Quality Appraisal

Mixed-Method Appraisal Tool (MMAT) (version 2018) was a promising and reliable tool for critically appraising and used to assess the strength of the evidence quality. Based on the number of criteria met for the respective study designs, the tool yielded scores of 20%, 40%, 60%, 80% and 100%. In this systematic review, the quality appraisal scores of the selected studies were counter-checked by both reviewers to enhance the quality assurance.

### Data Extraction

The resulting records from the screening process were saved in Endnote (X7) for the effective elimination of duplication. Data abstracting from the selected studies were collected in an excel spreadsheet. A data extraction form was preliminarily developed, piloted with at least six eligible studies and updated as a final data-entry template. Moreover, limitations of the study, recommendations and remarks on the study results were also documented in the data extracting excel sheet.

### Data Synthesis and Analysis

After the detailed construe, all selected articles were categorised into four groups such as studies related to environmental persistence, studies related to influencing factors on the persistence of viruses, studies related to disinfection methods, and cross-cutting studies that included more than one variable. After the data extraction and entry into the finalised excel template, the collected key data were arranged, coded and sorted out the same data points. Descriptive analysis was elected and manual synthesis was applied for the narrative presentation.

### Ethical Consideration

Since this systematic review involves none of the human subjects, there is a very low ethical issue for this study. However, the ethical form of a systematic review proposal was completed. Ethical clearance and approval were obtained from the Ethics Committee of the Institute for Health Research, the University of Bedfordshire on 27th June 2020. The reviewers were not intentionally misinterpreted the findings of the articles included in this review and presented the summary of retrieved information in the best interest of validating an accurate and transparent conclusion.

## Results

### Summary of Selection Process

The total number of 6184 titles and abstracts (3130 from reviewer-1 and 3054 from reviewer-2) were found at the first step of the screening process. 51 relevant published papers (30 from reviewer-1 and 25 from reviewer-2) were identified from other data sources as additional. Of these, 2850 studies were found as duplicates and 3001 studies were as irrelevant. After cleaning out duplicate and irrelevant studies, 333 full-text papers were found includable. Among 333 papers, 282 were ineligible for full-text reviewing due to some reasons (see in S2 Fig.), and finally, 51 papers remained for this present review. The characteristics of these included studies were presented in the table-2 in terms of frequency and percentage.

### Characteristics of Included Studies

All selected 51 studies were quantitative studies in which cross-sectional analytical studies were mostly contributed (n=24, 47%) and non-randomized controlled trials were second-most contributed for this review (n=22, 43%). The others were longitudinal cohort studies (n=2, 4%), quantitative non-randomized studies (n=1, 2%), cross-sectional descriptive studies (n=1, 2%) and comparative case-control studies (n=1, 2%). The included studies were published in January 2020 and originated from 16 countries. Most studies included were from the USA (n=16, 31%) and China (n=12, 23%). When the brief review was taken on the studied area by countries, studies in Australia (n=2) focused on the persistence of SARS-CoV-2 in wastewater and the impact of temperature on SARS-CoV-2 in both clinical and non-clinical contexts. All Studies from China (n=12) mainly focused on the persistence of SARS-CoV-2 in environmental matrices (surfaces, faeces, air, wastewater and surface) in both clinical and non-clinical areas. Studies from France (n=2) were conducted for SARS-CoV-2 on environmental surfaces in both clinical and non-clinical areas. One study from Hong Kong explored SARS-CoV-2 on surfaces and in the air of hospital environments (Table 2).

**Table.2.**
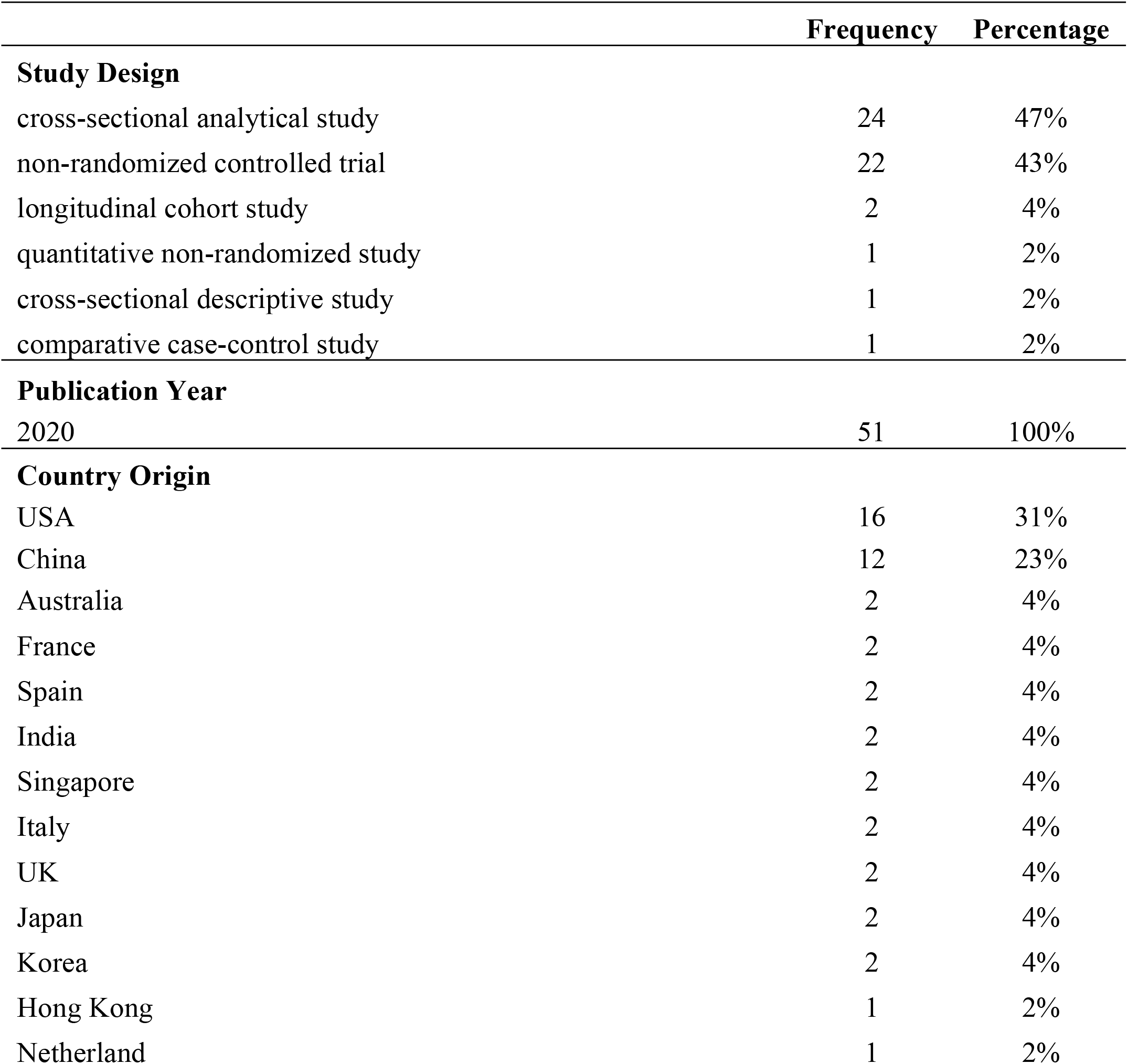
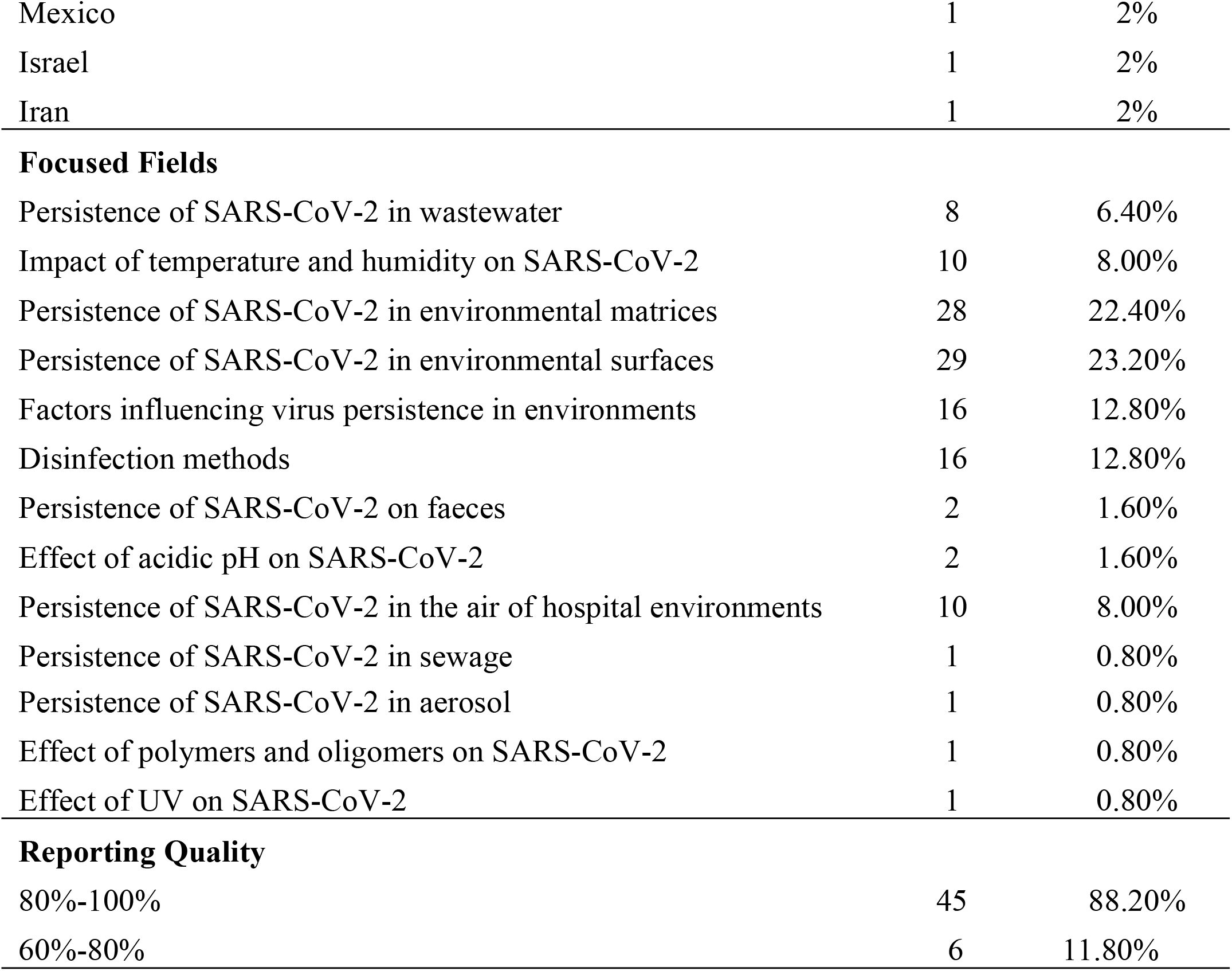
Characteristics of the Included Studies.

Two Indian studies emphasized SARS-CoV-2 RNA detection in wastewater. An Iran revealed SARS-CoV-2 RNA in air samples of hospitals. Two studies from Italy and two from the UK detected SARS-CoV-2 RNA in air, surfaces and wastewater while an Israel study was done on sewage measurements for SARS-CoV-2. Two papers from Korea studied environment contamination of SARS-CoV-2 on faeces and surfaces whereas 2 articles from Japan revealed surface and air contamination with SARS-CoV-2 and the effect of acidic pH on SARS-CoV-2. While a Netherland study measured SARS-CoV-2 aerosol, one study in Mexico explored the effectiveness of polymers and oligomers on the inactivation of the virus. Two studies in Singapore tested air and surface of clinical places for persistence of SAR-CoV-2 and a Spain study used UVG irradiator to examine the effects of UV on the virus. Sixteen studies from the USA covered all topics of environmental persistence of the virus, factors influencing virus persistence in environments and disinfection methods.

After appraising the methodological qualities of the selected studies using MMAT (Version 2018), papers in this review had a quality score ranging from 60% to 100%. Of 51 selected studies, 45 (88.2%) was occurred MMAT score of high quality (80-100 %) because those papers mentioned the specific description of in respective studies for the appropriate research problem approach, proper selection of target population and sampling methodology including consideration of confounding factors and appropriate measurements, appropriate explanation of intervention & exposure, completed outcomes summary with suitable analysis and well link between results and interpretation. Six studies (11.8%) had scored 60-80% for quality appraisal. The most distinct reasons for reduced scores were the unclear selection of the target population of the study, missed consideration of confounders and weak linkage or explanation between results and interpretation.

### Persistence of SARS-CoV-2 in Environmental Matrices/ Surfaces

Apropos of environmental matrices, not only on fomite surfaces and air media but also faeces and wastewater become concerned. Table 3 demonstrated the persistent durations of SARS-CoV-2 RNA in different environmental matrices/surfaces.

**Table.3.**
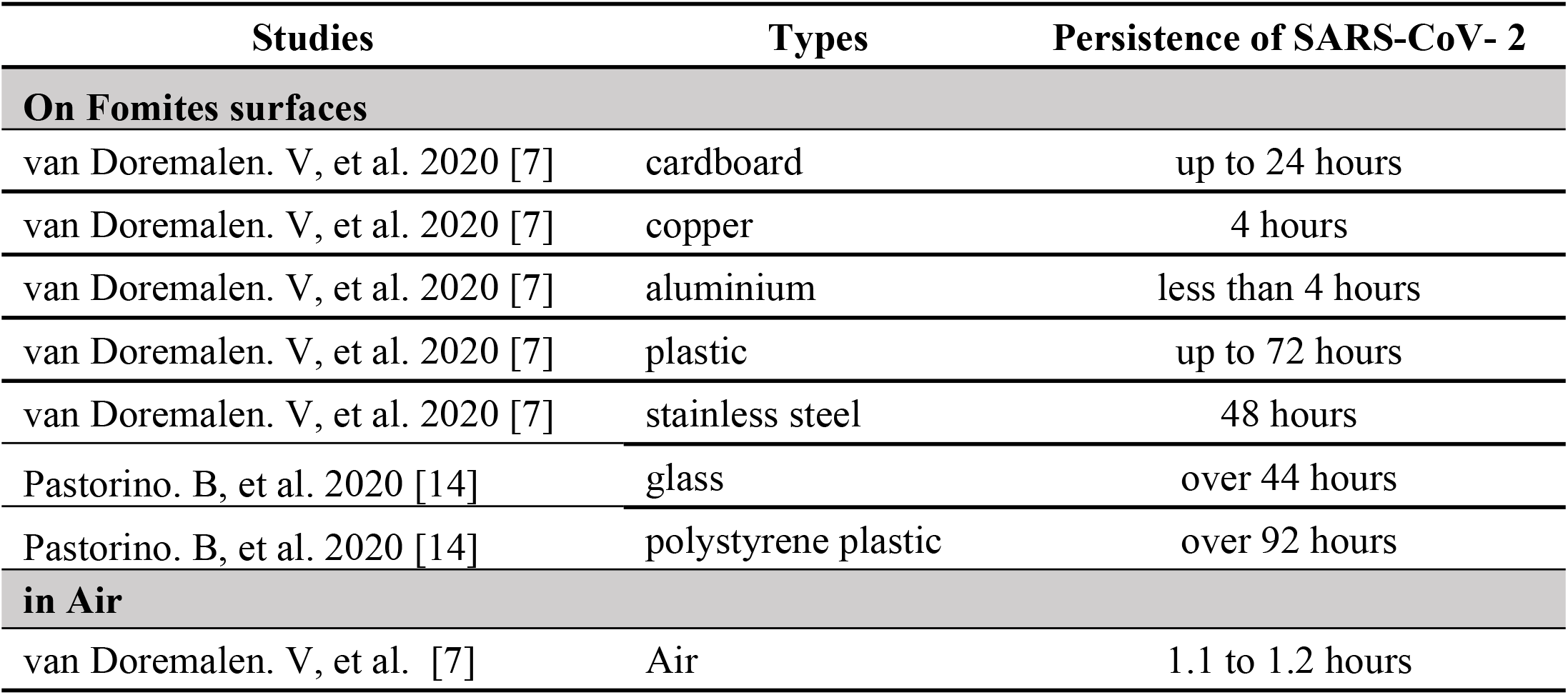
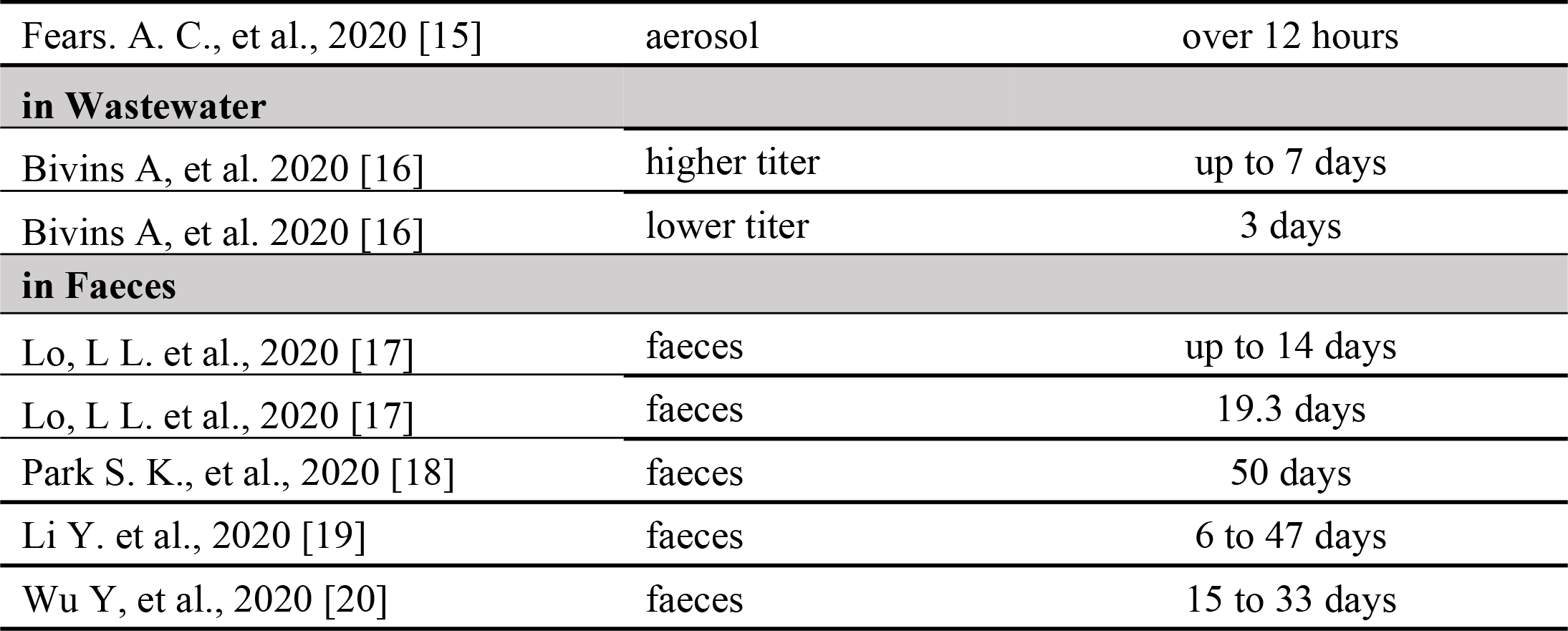
Persistence Durations of SARS-CoV-2 (Covid-19) in Environmental Matrices/Surfaces.

### Influencing factors for SARS-CoV-2 persistence in environments

#### Temperature and Relative Humidity (RH)

Effect on half-lives of SARS-CoV-2 varied with different combinations of temperature and RH. Virus half-lives were prolonged for 27 hours at 10 °C with 40% RH and were reduced to one and half hours at 27 °C and 65% RH. The estimated mean half-lives of the virus were 15.33 ± 2.75 hours with 20% RH, 11.52 ± 1.72 hours with 40% RH, 9.15 ± 3.39 hours with 60% RH and 8.33 ± 1.80 hours with 80% RH at 24°C respectively. The mean half-lives of the virus were estimated as 7.33 ± 1.33 hours with 20% RH, 7.52 ± 1.22 hours with 40% RH and 2.26 ± 1.42 hours with 60% RH at 35°C. Also, virus-like particles (VLP) survived better when it was incubated at 22°C than that at 34°C under dry conditions [15, 21]. The persistent durations of SARS-CoV-2 (Covid-19) at different temperatures were presented in Table 4.

**Table.4.**
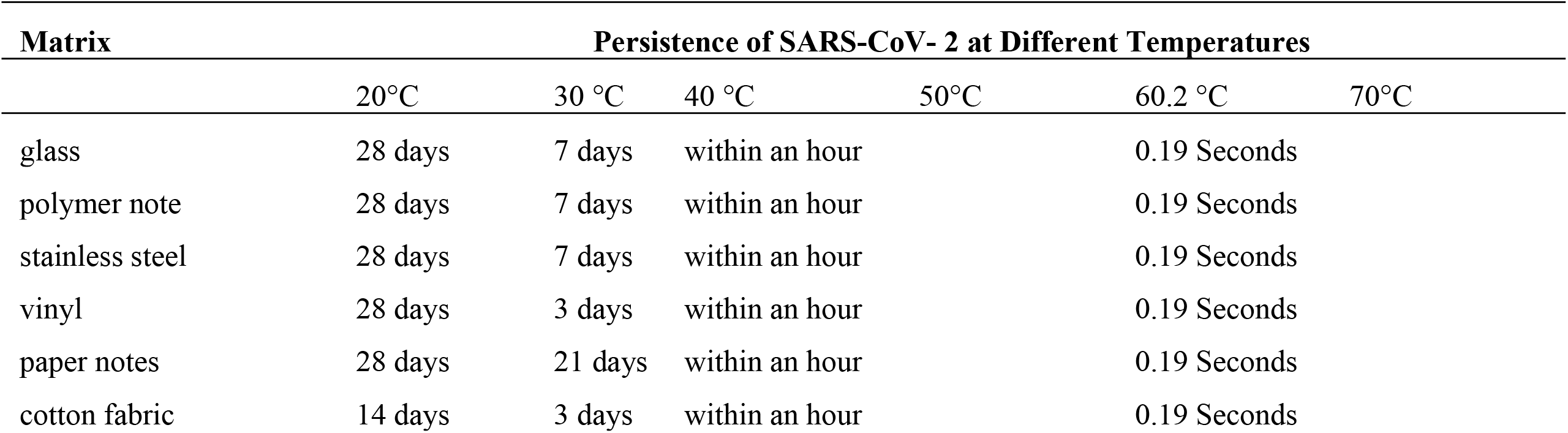
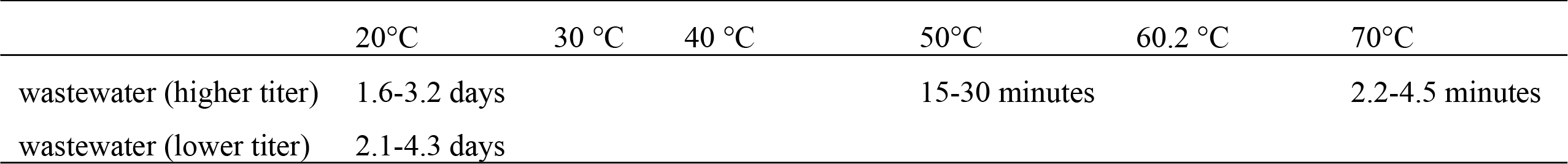
Persistence of SARS-CoV-2 at Different Temperatures.

| Matrix | Persistence of SARS-CoV- 2 at Different Temperatures |  |  |  |  |  |
| --- | --- | --- | --- | --- | --- | --- |
|  | 20°C | 30 °C | 40 °C | 50°C | 60.2 °C | 70°C |
| glass | 28 days | 7 days | within an hour |  | 0.19 Seconds |  |
| polymer note | 28 days | 7 days | within an hour |  | 0.19 Seconds |  |
| stainless steel | 28 days | 7 days | within an hour |  | 0.19 Seconds |  |
| vinyl | 28 days | 3 days | within an hour |  | 0.19 Seconds |  |
| paper notes | 28 days | 21 days | within an hour |  | 0.19 Seconds |  |
| cotton fabric | 14 days | 3 days | within an hour |  | 0.19 Seconds |  |
|  | 20°C | 30 °C | 40 °C | 50°C | 60.2 °C | 70°C |
| wastewater (higher titer) | 1.6-3.2 days |  |  | 15-30 minutes |  | 2.2-4.5 minutes |
| wastewater (lower titer) | 2.1-4.3 days |  |  |  |  |  |

#### Sunlight/ UV

Virus infectivity was reduced by far-UVC (222 nm) light for 90% in 8 minutes, 95% in 11 minutes, 99% in 16 minutes and 99.99% in 25 minutes with the dose of 1.2 mJ/cm^2^ to 1.7 mJ/cm^2^ [22].

#### pH/ Acidity

The pH 2.5 with free available chlorine (FAC) was identified as a potent deactivator for SARS-CoV-2 signifying >99.99% reduction of virus infectivity. Technically, the test solution, acidic electrolyzed water (EW), and ratio played a vital role in the inactivation process. By using the acidic EWs (pH-2.5, FAC-74 ppm) with a 1:9 ratio of the virus: acidic EW, virus titer was reduced by ≥4.25 log_10_ TCID_50_/mL with ≥99.99% reduction of infectivity after a one-minute reaction. However, neither visible reduction of virus infectivity could be identified on testing with a 1:1 ratio, an equal volume of virus and acidic EW. Moreover, a 17-days old solution of acidic EW (pH-2.5, FAC-109 ppm) yielded inferior action on deactivation of the virus compared to that of fresh acidic EW solution and 31-day stored acidic EW showed no detectable reduction in the virus infectivity [23].

#### Disinfection Methods in Healthcare Setting

With quaternary ammonium-based detergent for the floor, a sodium hypochlorite for non-floor surfaces in inpatient rooms and hydrogen peroxide for areas outside the patient rooms, surface disinfection was found effective in the reduction of SARS-CoV-2 persistence. As evidence, 36% of positive samples were reduced to 20% after cleaning with the above-mentioned disinfectants. When the disinfecting process was performed more frequently and thoroughly on floors with 2,500 ppm sodium hypochlorite, the persistence of the study virus in surface samples was found significantly reduced by 3.4% [24]. After cleaning with the combined disinfectant and detergent (e.g., Surfanios Premium), the contamination in samples was considerably reduced from 60% to 4.9% for the floors and all the surfaces directly in contact with patients (such as trolleys, skechers, cuffs, door handles etc.) and from 10% to 5.6% for the surfaces not directly in contact with patients (such as stethoscopes) [25]. Moreover, Pulsed Xenon Ultraviolet (PX-UV) reduced the infectivity of SARS-CoV-2 to 99.97% at 1 minute, 99.997% at 2 minutes and 99.992% at 5 minutes. Contamination of N95 respirators in inoculation was reduced to 99.998% with 2 minutes of exposure with PX-UV [26].

For the effective decontamination of respirators, the recommended dose and time needed to expose to UV light were 5 mj/cm^2^ UV dose for 11 seconds, 300 mj/cm^2^ for 12 minutes, 1 j/cm^2^ for 36 minutes and 3 j/cm^2^ for 1 hour 40 minutes [27]. For filtering faceplate respirators (FFRs), a dose of 1 mj/cm^2^ of UV-C was a bottom need for disinfecting. Biosafety cabinets (BSC) were used for minimum level UV irradiation to achieve the target dose of decontamination on FFRs. The minimum duration of irradiation for FFRs was identified as 4.3 hours per side, for PPE as 62 minutes per side and for face shields as 15.6 minutes per side (60 mj/cm^2^ of UV radiation) [28]. Furthermore, decontamination of SARS-CoV-2 on 3M-N95 with UVC in germicidal UVC device was also discussed that total disinfection was attained within 120 seconds. 1 log reduction of viral titer was identified in 2 seconds of UV exposure per side and 2 log in 54 - 120 seconds per side [29]. Regarding decontamination of SARS-CoV-2 in wastewater, preliminary disinfection in septic tanks was performed with free chlorine N6.5 mg/L for 1.5 hours with the dosage of sodium hypochlorite (800 g/m^3^). However, 12 hours after sodium hypochlorite had been added to septic tanks, the study virus RNA was significantly detectable again due to the decline of free chlorine. When sodium hypochlorite was increased to 6700 g/m^3^, SARS-CoV-2 became undetectable in wastewater [30].

#### Disinfection Methods in Non-healthcare/General Setting

With the oligomers disinfectants activated by UV light, complete disinfection happened within 10-15 minutes. However, the effectiveness of oligomers became lower in dark places than that occurred under the light [31]. SARS-CoV-2 infectivity was reduced to >90% in 10 minutes and >99.99 % in 2 hours on the antimicrobial treated surfaces of stainless steel [32]. Regarding indoor environments, Far UVC light (222 nm) showed its effectiveness to deactivate the SARS-CoV-2 as 90% in 8 minutes, 95% in 11 minutes, 99% in 16 minutes and 99.9 % in 25 minutes [22]. UVB irradiation at 1.6-0.7 W/m^2^ also deactivated the SARS-CoV-2 faster than that at 0.3 W/m^2^. The effective decontamination of the SARS-CoV-2 in wastewater was achieved by the wastewater treatment, particularly including secondary treatment-(Moving Bed Biofilm Reactor (MBBR), Sequencing Batch Reactor (SBF) and Activated Sludge Process (ASP)) and tertiary treatment-(chlorine and UV) [33].

## Discussion

Until the end of the paper screening process i.e., October 2020, by the Author’s knowledge, this systematic review was the only comprehensive review covering the three areas of environmental persistence of SARS-CoV-2 in different matrices, influencing factors of the virus persistence and disinfection methods obtained from the primary studies full-text published from January 2020. Moreover, this review exclusively focused on the SARS-CoV-2 virus and explicitly included both healthcare settings and non-healthcare settings.

### SARS-CoV-2 Virus Persistence in Environmental Matrices/ Surfaces

Based on the findings of the review, the possible persistence of SARS-CoV-2 in environmental matrices was <4 hours on aluminium, 4 hours on copper, 24 hours on cardboard, 44 hours on glass, 48 hours on stainless steel, 72 hours on plastic and 92 hours on polystyrene plastic. These findings had some variations compared with the findings of Kampf, et al.’s review documented the survival of the coronavirus family on aluminium for 2-8 hours, latex rubber for ≤8 hours, glass for 4 days, plastic for 2-6 days, steel/silicon rubber/ceramic/Teflon for 5 days, a disposable gown for 2 days, metal for 5 days, wood for 4 days and paper for 4-5 days [34]. In this review, the most contaminated objects with SARS-CoV-2 in inpatient rooms and staff areas were the mobile phones of the patients, buttons of water machines, elevators, beepers, doorknobs and hand sanitiser dispensers, printers, desktops, keyboards and eye protection/face shields/gloves. In non-healthcare areas, the SARS-CoV-2 virus was detected in the air with an approximate half-life of 1.1-1.2 hours and even robust over 12 hours in aerosol form. The common areas with virus aerosol were found not only in the mobilised areas of Covid-19 patients but also in general public areas and general wards of the hospitals. These findings were consistent with the findings of Tang et al.’s review in which the infectivity of SARS-CoV-2 aerosols was identified as extending up to 16 hours. The risk of SARS-CoV-2 aerosol was also classified as high risk (for healthcare settings/ laboratory) and medium to low-medium risk (for public transportation/ naval vessels, public places, restrooms, churches, prisons, schools, nursing homes, and kindergarten areas) in Tang et al.’s review [35]. In this review, contaminated wastewater with SARS-CoV-2 could be infectious for 3-7 days. To compare, Rosa *et al*.’s review stated that the SARS-CoV virus persisted in wastewater for 2 days at 20 °C and ≥ 14 days at 4°C [36]. This review concluded that the duration of SARS-CoV-2 persistence in faeces was 14-50 days and remained positive up to 15-33 days with negative respiratory samples. The review of Gupta *et al*. mentioned the same outline for the faecal contamination that was positive for 3-30 days from the onset of symptoms and 3-21 days after the negative nasopharyngeal test [37].

### Influencing factors for SARS-CoV-2 persistence in environments

By changing temperature and humidity, SARS-CoV-2 persistence in environments fluctuated. SARS-CoV-2 persistence on non-porous and porous surfaces was recorded as 28 days and 14 days at 20°C and 7 days and 3 days at 30°C respectively. At 40 °C, 99.99% reduction of virus infectivity on all fomite surfaces within an hour. However, the virus could survive up to 21 days on paper notes at 30°C. Kampf. et al.’s review, though, reported approximately 5 days for non-porous surfaces at room temperature or 20°C [34]. This systematic review identified additional facts on the impact of temperature on viral load reduction and the effect of relative humidity (RH) on SARS-CoV-2 persistence in environments which was not included in Kampf, et al [34]. This particular review identified that virus decaying was noted approximately 5-10 times faster at 27°C than that at 10°C and 2-5 times even faster with 65% RH than that with 40% and 100% RH. Quick virus decaying was recognized with high temperature and RH (35°C with 60% RH). The virus half-life varied with changing temperature and RH such as 15.33±2.75 hours at 24°C with 20%RH and 2.26±1.42 hours at 35°C with 60%RH indicating the virus was mostly stable at ambient indoor temperature with relatively low RH. This review also identified that >3-4 days at 20°C, 30 minutes at 50°C and only over 4 minutes at 70°C were needed for 99 % reduction of the SARS-CoV-2 infectivity in wastewater. The results denoted that the higher titer virus stayed more days in wastewater than the low titer at the same temperature. The higher the temperature was, the faster the reduction of the SARS-CoV-2 infectivity occurred in wastewater. This review’s findings on temperature’s impacts on the virus infectivity were consistent with the findings from Rosa, et al. ‘s review where SARS-CoV could survive ≥14 days at 4°C and 2 days at 20°C. This review also concluded that the virus infectivity was reduced by 99.99% in 25 minutes by far-UVC (222 nm) light with a dose of 1.2 mj/cm^2^ to 1.7 mj/cm [2, 36]. This finding was in line with the findings of Riddell et al.’s review described that the virus was susceptible to the UV light around 253.7 nm. This review also identified the critical role of free available chlorine (FAC) concentration in acidic EW activities on the reduction of SARS-CoV-2 infectivity and recorded that acidic EW (pH-2.5), FAC-74 ppm, potently reduced >99.99% of SARS-CoV-2 infectivity with a 1:9 ratio of virus: acidic EW solution [38]. In contrast, Cervino et al.’s review recorded that the range of pH (3-10) did not show any significant changes in the stability of SARS-CoV-2 [39].

### Disinfection Methods

Based on the review findings, ammonium-based detergent, sodium hypochlorite, hydrogen peroxide and combined disinfectant and detergent were recommended for effective decontamination on surfaces and floors. Besides, polymers and oligomers were identified as significant disinfectants for SARS-CoV-2. These agents effectively deactivated the virus under UV light within 10-15 minutes. However, the deactivation actions of these agents did not occur in the darkness. Antimicrobial treatment on the stainless steel was noted for the reduction of SARS-CoV-2 infectivity to >99.99 % in 2 hours. The mentioned findings agreed with Kampf et al. in which a range of disinfectants was recorded as effective decontamination for coronavirus by 4 log_10_ with 78%-95% ethanol, 70%–100% 2 propanols, combined of 45% 2 propanols with 30% 1 propanol, 0.5%-2.5 % glutardialdehyde, 0.7%-1% formaldehyde, 0.23%–7.5% povidone-iodine, at least 0.21% sodium hypochlorite and 0.5% hydrogen peroxide in an exposure time of 15 seconds to 10 minutes approximately [35]. In this review, far UVC light (222 nm), UVB irradiation (1.6 - 0.7 W/m^2^), and Pulsed Xenon Ultraviolet (PX-UV) (200–320 nm) deactivated SARS-CoV-2 in indoor environments. This review revealed that contaminated N95 respirators were disinfected 99.998% at 2 minutes of exposure with PX-UV in the UVGI box. Notably, UV light had the effective decontamination of respirators, medical equipment and PPE, however, the benefit of utilisation should be outweighed by the harmful effect and cost. Regarding decontamination of SARS-CoV-2 in wastewater, sodium hypochlorite (6700 g/m3) with free chlorine was effective for preliminary disinfection in septic tanks. Compared with Carraturo et al.’s review in which sodium hypochlorite with 10 mg/l dosage provided an effective reduction of infectivity to 5 logs in 30 minutes of exposure [40]. However, it would be quite concerned to add the recommended dose of sodium hypochlorite per litre of wastewater to maintain decontamination if the wastewater volume was plenteous. Furthermore, MBBR (Moving Bed Biofilm Reactor), SBF (Sequencing Batch Reactor) and ASP (Activated Sludge Process) were recommended as the effective disinfectants in the secondary treatment of wastewater. For the tertiary treatment of wastewater, chlorine and UV were recommended as effective. This systematic review explored the potential spreading sources of COVID-19 in contaminated areas and the influencing factors for the virus in environments since these could be key determinants for the prolongation of the COVID-19 outbreak. Last but not the least, this research revealed effective disinfection methods to break the chain of transmission of SARS-CoV-2 to prevent the COVID-19 surge.

## Conclusion

Since the COVID-19 pandemic has been pestilential over a considerable duration, global deployment and financial crisis could not be reversed as before. It brought up essentials to allow the nations fully back to work with effective preventive measures. This systematic review documented the key findings that came out through the hard work of the 51 studies across 16 countries. The findings of this systematic review reflected a comprehensive overview of the persistence of SARS-CoV-2 in different environmental matrices under different conditions and the effects of disinfectants and their techniques on the viability of SARS-CoV-2 in both healthcare and non-healthcare settings. Those findings will be important inputs for authorities in the development of mitigation strategies and policies for effective preventive measures for COVID-19. This particular review advocated that the persistence of SARS-CoV-2 in environments should be counted in consideration of disinfection methods and materials. Besides, the infection control team should be informed to develop proper disinfection guidelines/instructions based on the evidence of review. To optimise, the cleaning interval, mechanism and the agents/methods used for decontamination should be regularly monitored with sampling and testing by a defined supervision team. In conclusion, additional research on the possible food contamination, the weather/climate effects and fumigation/spraying effects on the virus should be advanced.

## Limitations of Study

While this study was focusing to review the valuable evidences systematically with the predefined criteria, there were some limitations. The exclusive inclusion of the articles which was published in full-text could result in missing of some information from unpublished studies. Besides, this review could not reveal some articles that might exit in other languages rather than English. Since the review was dedicated to the academic purpose, there was a time constraint leading to the paper selection process needed to finalize in October 2020 so that published paper later than October 2020 could not be included in this review. Moreover, this review did not include the discussion on the possible food contamination, the weather/climate effects and fumigation/spraying effects on the virus. With the above-mentioned limitations, the information and discussion conveyed in this systematic review may be arguable for certain extent on making a definite conclusion of SARS-CoV-2 persistence in environmental matrices, influencing factors and disinfection methods.

## Access to data

Details of authors’ contacts including phone numbers, emails, and affiliations were mentioned on the cover page of the manuscript for being requested additional information such as the extracted data sheets and tables.

## Dissemination plan

This dissertation was documented at both of the University of Bedfordshire, UK and the STIMU, Myanmar as a part of the academic procedure. A short report of this review would be circulated among medical and public health professionals in Myanmar. Since the study area in this dissertation focused on the current concern of the global health crisis and also registration process was done through PROSPERO (International Prospective Register of Systematic Reviews) on 13th October 2020 as a part of this systematic review, it is expected to be published in the PLOS One, a peer-reviewed open access scientific journal.

## Supporting information

Supplemental Sheets 1

Supplemental Sheet 2

## Administrative information

The authors declare that there was no conflict of interest and they have adhered to all principles of academic honesty and integrity and have not misrepresented any data or information intentionally in this report. This report is a part of the dissertation submission that was dedicated to the University of Bedfordshire for the gratification of a Master of Science in Public Health.

## Author contributions

Research question: Chaw Chaw Yu

Research Design: Chaw Chaw Yu

Research protocol: Chaw Chaw Yu, Thein Hlaing, Kyaw Myo Tun

Searching Literature: Chaw Chaw Yu, Thein Hlaing

Selection of Studies: Chaw Chaw Yu, Thein Hlaing

Critical Appraisal: Chaw Chaw Yu, Thein Hlaing Data Extraction: Chaw Chaw Yu,

Data Synthesis and Analysis: Chaw Chaw Yu Writing: Chaw Chaw Yu

Writing (revision & editing): Chaw Chaw Yu, Kyaw Myo Tun

## Acknowledgements

The authors would like to express sincere thanks to the research supervisor, co-reviewers, course lecturers from both Universities (STIMU and the University of Bedfordshire, UK) and batch-mates for their academic support, kind guidance and encouragement to conduct this dissertation on systematic review and for successful submission on schedule.

## Supporting information

S1 Table. PRISMA (Preferred Reporting Items for Systematic Review and Meta-analysis) 2020 checklist

S2 Figure. Flow Diagram of Study Selection Process

---

**Figure 1:**
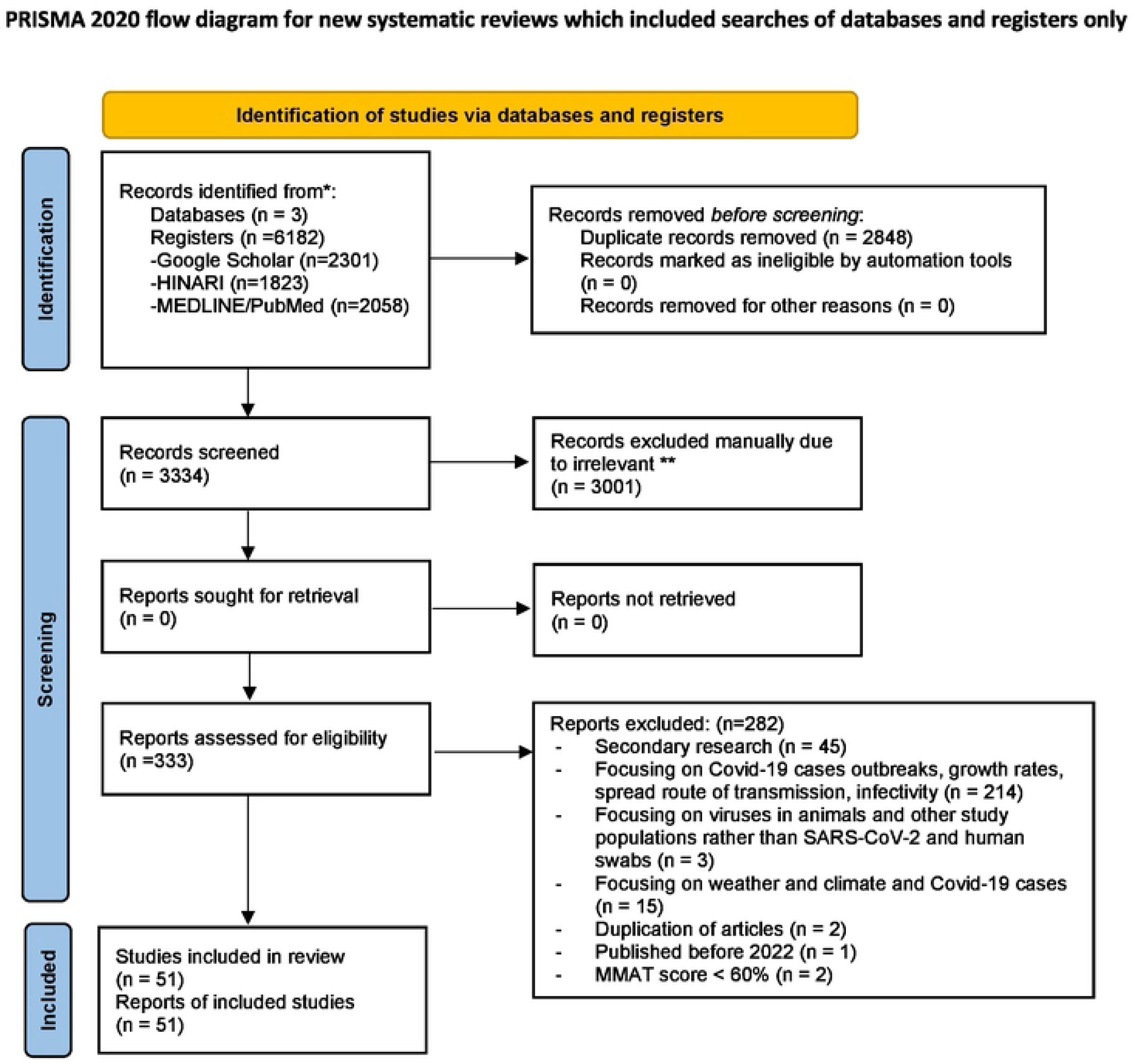
Included/Excluded Literature * Consider, if feasible to do so, reporting the number of records identified from each dalabase or register searched (rather than the total number across all databases/registers). ** If automation tools were used, indicate how many records were excluded by a human and how many were excluded by automation tools. *From*: Page MJ, McKenzie JE, Bossuyt PM, Boutron I, Hoffmann TC, Mulrow CD, et al. The PRISMA 2020 statement: an updated guideline for reporting systematic reviews. BMJ 2021;372:n71. doi: 10.1136/bmj.n71 For more information, visit: http://www.prisma-statement.onq/

## Notes

### Competing Interest Statement

The authors have declared no competing interest.

