## Supplemental Sheet 2 for "Influencing Factors for the Persistence of SARS-Cov-2 (Covid-19) exposed in Environmental Matrices and Disinfection Methods: Systematic Review"

**Supplemental Sheets for Methodology: Detailed Description for Bibliographic Search**

The prime databases used for searching articles were Google Scholar, HINARI and MEDLINE/PubMed. Other databases such as EBSCOHOST and Cochrane Library were also explored. By using the predefined keywords, pilot screening was performed and keywords were modified accordingly. Besides, the search strategy was tested with Boolean Operators by two independent investigators and compared the total number of eligible papers found. Whenever significant differences in the number of searched papers had occurred, thorough discussions between investigators were performed to optimise the keywords and searching mechanism. The potential title and abstract screening process was initiated after the confirmation of keywords and search strategy. The full-text screening was then proceeded to filter the eligible articles. Additionally, a manual search and screening of the reference lists were elaborated. Moreover, Medical Subject Headings (MeSH) terms and specific symbols such as the asterisk-(*) marks and dollar signs-($) were combined in the search mechanism to identify truncation or find the related terms to find relevant articles.

Keywords identified for search process are listed as in Table (1).

**Table (1) Keywords identified for Search**

| Interest problem  Concept (I) | Intervention  Concept (II) | Outcome  Concept (III) | Others  Concept (IV) |
| --- | --- | --- | --- |
| SARS-CoV-2  Covid-19  Human Coronavirus | Exposed  Exposure  Contact | Persistence  Survival  Live  Factors influencing  Environments  Environmental matrices  Surfaces  Objects  Disinfection  Decontamination  Methods | Healthcare setting  Clinical setting  Non-healthcare setting  Household  Common public place  Public space |

**Table (2) Search Strategy Using Boolean Operators**

| Problem  Concept (I) | AND/  OR | Intervention  Concept (II) | AND/  OR | Outcome  Concept (III) | AND/  OR | Others  Concept (IV) |
| --- | --- | --- | --- | --- | --- | --- |
| SARS-CoV-2  **OR**  Covid-19  **OR**  Human Coronavirus |  | Exposed  **OR**  Exposure  **OR**  Contact |  | Persistence  **OR**  Survival  **OR**  Live  **OR**  Factors influencing  **OR**  Environments  **OR**  Environmental matrices  **OR**  Surfaces  **OR**  Objects  **OR**  Disinfection  **OR**  Decontamination  **OR**  Methods |  | Healthcare setting  **OR**  Clinical setting  **OR**  Non-healthcare setting  **OR**  Household  **OR**  Common public place  **OR**  Public space |

By Boolean Operators, which was used to search the relevant articles for review process to answer the predefined research question, each keyword was searched as free text term search. Then, All keywords was combined with OR, followed by each concept will be combined with AND.

Detailed description of search items with Boolean Operators was stated as follows.

a) For concept (I), two steps were performed for search. At first step, each keyword listed in table-1 was searched separately such as; SARS-CoV-2, Covid-19, Human Coronavirus. As second step, all keywords in concept (I) (listed in table-1) were combined with OR such as; SARS-CoV-2 OR Covid-2 OR Human Coronavirus.

**Table (3) Example of Boolean Operators Search Strategy in Details**

| For Concept (I) | |
| --- | --- |
| First step | **Second step** |
| SARS-CoV-2  Covid-19  Human Coronavirus | SARS-CoV-2 OR Covid-2 OR Human Coronavirus |

b) Two steps performed in concept (I) had to be repeated for the other concept of (II), (III), (IV) from table (2) as shown in table (3).
c) Then, OR search (i.e. second step) of all concept was combined with AND such as second step of concept (I) AND second step of concept (II) AND second step of concept (III) AND second step of concept (IV)
d) OR search (i.e. second step) of 3 concepts was combined with AND such as second step of concept (I) AND second step of concept (II) AND second step of concept (III).
e) OR search (i.e. second step) of 3 concepts was combined with AND such as second step of concept (I) AND second step of concept (II) AND second step of concept (IV).
f) OR search (i.e. second step) of 3 concepts was combined with AND such as second step of concept (I) AND second step of concept (III) AND second step of concept (IV).
g) OR search (i.e. second step) of 2 concepts was combined with AND such as second step of concept (I) AND second step of concept (II)
h) OR search (i.e. second step) of 2 concepts was combined with AND such as second step of concept (I) AND second step of concept (III)
i) OR search (i.e. second step) of 2 concepts was combined with AND such as second step of concept (I) AND second step of concept (IV)

Apart from the simple combination of search by Boolean Operators, complex combination was also used for vigorous search such as

a) [second step of concept (I) AND second step of concept (II) ] AND [second step of concept (III) OR second step of concept (IV)]
b) [second step of concept (I) AND second step of concept (IV) ] AND [second step of concept (II) OR second step of concept (III)]
c) [OR search (i.e. second step) of 3 concepts will be combined with AND such as second step of concept (I) AND second step of concept (II) AND second step of concept (III)] OR second step of concept (IV)
d) [OR search (i.e. second step) of 3 concepts will be combined with AND such as second step of concept (I) AND second step of concept (III) AND second step of concept (IV)] OR second step of concept (II)
e) [OR search (i.e. second step) of 3 concepts will be combined with AND such as second step of concept (I) AND second step of concept (II) AND second step of concept (IV)] OR second step of concept (III)
f) Second step of concept (I) AND [second step of concept (II) OR second step of concept (III)]
g) [second step of concept (I) AND second step of concept (II) ] OR second step of concept (III)
h) [second step of concept (I) AND second step of concept (III) ] OR second step of concept (II)

Throughout the searching process, two reviewers worked independently for search but cooperatively resolved if there was any disagreement on search results. Call for third party (herein supervisor) was involved for any consensus necessities.
